# Deep Clinical Phenotyping of Planar Perugini Grade 1 on Bone Scintigraphy: An Imaging-Phenotype Association Study

**DOI:** 10.64898/2026.06.29.26356893

**Authors:** Clemens P. Spielvogel, Jing Ning, Katharina Mascherbauer, Katarina Kumpf, Michael Poledniczek, David Kersting, René Rettl, Felix Hofer, Christian Hengstenberg, Marcus Hacker, Christian Nitsche, Raffaella Calabretta

**Author notes:** Correspondence to: Clemens Spielvogel, PhD Division of Nuclear Medicine, Department of Biomedical Imaging and Image-guided Therapy Medical University of Vienna Spitalgasse 23, 1090 Vienna, Austria ORCID: 0000-0002-4409-8324. Christian Nitsche and Raffaella Calabretta share the last authorship.

## Abstract

**Aims:** Planar Perugini grade 1 is equivocal for transthyretin amyloid cardiomyopathy (ATTR-CM), has no dedicated management pathway and is heterogeneous, spanning low-burden amyloid and non-amyloid blood-pool activity. Rather than treating it as a step toward amyloid confirmation, we characterized the clinical phenotype grade 1 marks and tested whether that phenotype accounts for its prognosis.

**Methods and Results:** We studied 9,170 consecutive patients undergoing [^99m^Tc]Tc-DPD bone scintigraphy. Grade was assigned by blinded expert consensus on planar images, per the original Perugini definition. Using an imaging-phenotype association (IPA) framework, we linked grade to 1,243 clinical, laboratory, echocardiographic, CMR and ICD-10 parameters. The endpoint was a composite of heart failure or death. Grade 1 occurred in 175 (1.9%) and grade ≥2 in 142 (1.6%) patients. Grade ≥2 reproduced the infiltrative ATTR phenotype, with greater septal thickness, higher extracellular volume, neuropathy and atrial fibrillation, validating the framework. Grade 1 instead marked a non-infiltrative cardiometabolic phenotype dominated by hypertension, chronic ischemic heart disease, high BMI, anemia, reduced eGFR and atrial enlargement. Low extracellular volume and septal thickness argued against infiltration. Grade 1 carried worse outcomes than grade 0 (adjHR 1.30, 95% CI 1.07-1.59). Within grade 1, a phenotype of hypertension, high BMI and low hematocrit reached or exceeded grade ≥2 risk (HR 2.46 vs 1.76).

**Conclusion:** Planar grade 1 is neither uniform early amyloid nor a benign artifact but a mixed-etiology, predominantly non-infiltrative cardiometabolic phenotype carrying clinically meaningful risk. This supports reinterpreting grade 1 and prioritizing cardiorenal comorbidity alongside selective amyloid workup.

## Introduction

Cardiac amyloidosis is an infiltrative cardiomyopathy caused most often by transthyretin (ATTR) or immunoglobulin light chain (AL) deposition. Bone-seeking [^99m^Tc]-labeled diphosphonates are central to its noninvasive evaluation and planar bone scintigraphy remains among the most frequently performed nuclear imaging studies worldwide owing to its availability, low cost and established diagnostic value (1–3). Moderate to strong cardiac uptake (Perugini grade ≥2) in the absence of a monoclonal protein is widely accepted as diagnostic for ATTR-CM without biopsy (4,5).

Grade 1 is different. Mild uptake is equivocal, does not establish a diagnosis and is currently not assigned a dedicated management pathway. Mechanistically, it is heterogeneous. When confirmed to present myocardial uptake on SPECT, it may reflect early or low-burden ATTR or AL, yet SPECT/CT studies show that in most grade 1 scans, planar signal reflects blood-pool rather than myocardial activity (6–9). At the same time, planar grade 1 is not prognostically irrelevant. All-comer analyses report worse outcomes in grade 1 than in patients without uptake (10,11). These observations sit in tension. If most grade 1 is not myocardial amyloid, an association with adverse outcomes seems counterintuitive.

The prevailing framing treats grade 1 as a faint or early amyloid signal to be confirmed or excluded. We assessed the opposite view, that the prognostic value of grade 1 may lie in the clinical company it keeps rather than in amyloid itself. Resolving this requires moving beyond small confirmatory SPECT series toward a systematic, high-dimensional characterization of patients with grade 1 on planar scintigraphy.

We therefore applied an imaging-phenotype association (IPA) framework, analogous to genome-wide and phenome-wide association studies (GWAS), to a large consecutive all-comer cohort undergoing bone scintigraphy. By relating Perugini grade to 1,243 clinical, laboratory, imaging variables and diagnoses, we aimed first to validate the approach by recovering the established infiltrative phenotype of grade ≥2 and second, to characterize the clinical phenotype marked by grade 1. We then tested the central hypothesis that grade 1 prognosis is driven substantially by an underlying cardiometabolic phenotype by asking whether a phenotype defined on routine comorbidity and laboratory markers identifies grade 1 patients with increased risk.

## Material and methods

### Study design and participants

Patients consecutively referred for whole-body [^99m^Tc]Tc-DPD scintigraphy at the Medical University of Vienna (Division of Nuclear Medicine, Vienna, Austria) between January 2010 and July 2020 were included for analysis. Patient data were extracted from electronic health records, including demographic characteristics, blood biomarkers, echocardiographic findings, cardiac magnetic resonance (CMR) imaging parameters, comorbidities and [^99m^Tc]Tc-DPD scintigraphy. For patients undergoing multiple scintigraphic examinations, only the earliest available scan was used in this study to ensure statistical independence. In an exploratory subset of grade 1 patients, SPECT/CT was available and was used to categorize tracer localization as myocardial or blood-pool. Given the small size of this subset, these analyses were treated as hypothesis-generating.

The study had three objectives. First, to validate the IPA framework by recovering established associations between clinical parameters and amyloid-suggestive grade ≥2 uptake. Second, to characterize the clinical phenotype marked by grade 1. Third, to test whether grade 1 prognosis is driven by a cardiometabolic phenotype rather than by amyloid, by determining whether that phenotype identifies grade 1 patients whose risk approaches or exceeds that of grade ≥2.

The study was conducted in accordance with the Declaration of Helsinki and approved by the Institutional Review Board of the Medical University of Vienna (2278/2024). Owing to the retrospective design, the requirement for informed consent was waived.

### Imaging data acquisition and interpretation

Planar whole-body [^99m^Tc]Tc-DPD scintigraphy was performed according to established clinical practice guidelines, with imaging acquired ≥2 hours after tracer administration (12). Tracer uptake was categorized using the Perugini grading scale (13). Perugini grade was always assessed solely based on planar scintigraphy, regardless of SPECT imaging or other examination results. As patients with grades 2 and 3 have the same clinical interpretation, they were analyzed together (13). To ensure accurate grading and reduce inter-reader variability, image interpretation was conducted independently by three experienced readers (>5 years of experience) for the purpose of this study, blinded to clinical data and to one another’s assessments. In the event of discrepant readings, final grades were adjudicated by consensus with two additional imaging specialists, each with >10 years of experience. An additional three-dimensional SPECT/CT was performed after scintigraphy in a subset of patients ≥2 hours after tracer injection and blood pool versus myocardial uptake was evaluated by an experienced nuclear imaging expert (Raffaella Calabretta; 13 years of experience).

While current expert consensus recommends confirming positive Perugini grades on SPECT, the Perugini score is, by its original description, a visual scale applied to planar images (13). In this study, we did not differentiate blood pool from myocardial activity, because thoracic SPECT/CT was performed in only 24 of 175 (14%) grade 1 patients, largely predating that recommendation. All grades reported here are therefore planar grades, denoted planar grade 0, 1 and ≥2, and should not be equated with SPECT-confirmed grades. All studies were graded for the purpose of this analysis.

### Clinical data

Clinical parameters (n=55) included demographics (n=3), blood biomarkers (n=19), echocardiographic parameters (n=17) and CMR parameters (n=15), all extracted from electronic health records of the Vienna General Hospital (Vienna, Austria) for a period of up to six months before imaging. Documented comorbidities (n=1,189) were obtained from the admission and discharge reports from the Vienna Health Association (7 hospitals). Data were derived from routine clinical care EHRs and therefore contained missing values, which were imputed for downstream analyses using the median and mode for continuous and categorical data, respectively. Echocardiographic assessment of valvular function (tricuspid, pulmonary, mitral, aortic) included stenosis and regurgitation, dichotomized as moderate or greater versus less than moderate. Comorbidity data were derived from ICD-10 codes in EHR systems of the Vienna Health Association (7 Viennese hospitals). Other parameters were obtained from the EHR system of the Vienna General Hospital. For time-to-event analyses, a composite endpoint of heart failure hospitalization (HFH) or death was employed. Mortality data were obtained from the Austrian Death Registry and HFH events were identified from inpatient admission and discharge documentation of the Vienna Health Association, the Vienna General Hospital and the national electronic health record system. Missingness for clinical characteristics are reported in **Supplemental Table S1**.

### Imaging-phenotype association analysis

We adapted the association-mapping logic of genome-wide association studies to imaging, systematically relating each clinical trait to the imaging-derived Perugini grade with confounder correction (14). Traits comprised two classes, binary diagnoses from ICD-10 codes and continuous laboratory, echocardiographic and CMR measures. Variables with non-informative variance were removed. Continuous variables were median-imputed and binary variables mode-imputed. The Perugini grade was the independent variable and all models were adjusted for age, sex and past or present cancer, the last because of the high oncologic prevalence in this referral cohort.

Each trait was modeled separately, using logistic regression for binary traits and linear regression for continuous traits, both with heteroscedasticity-consistent standard errors. Binary traits required at least 10 positive cases to enter, avoiding sparse-data bias. Effects were reported as log2 odds ratios for binary traits and standardized coefficients for continuous traits, with Wald p-values. The grade ≥2 versus grade 0 contrast excluded grade 1 patients to keep the groups distinct. We controlled the family-wise error rate with Bonferroni correction, with the threshold set by the number of traits surviving filtering in each analysis and confirmed robustness with Benjamini-Hochberg FDR control at q<0.05. Associations are shown as volcano plots encoding direction, effect size, significance and sample-size weighting. Grade-specific prevalence for all ICD-10 traits is provided as a public supplementary resource.

### Cardiometabolic phenotype definition

To define a phenotype that distinguishes grade 1 from both neighboring grades, we filtered IPA-significant or amyloid-relevant parameters by an additional comparison of grade 1 to grade 0 and to grade ≥2. Binary parameters were compared with Fisher’s exact test and continuous parameters with the Mann-Whitney U test, using the risk difference and Cliff’s delta as effect sizes. From the joint result each parameter was labeled as resembling grade ≥2, resembling grade 0, distinct from both, or indeterminate. This step was descriptive and unadjusted for multiple comparisons.

We then constructed a cardiometabolic phenotype from parameters that distinguished grade 1 from grade 0 or grade ≥2, excluding parameters intermediate between the two grades to avoid capturing amyloid cases and excluding parameters available in under 50% of patients. The composite was fixed before any survival testing to limit optimization. We then employed the resulting, filtered parameters to define a primary phenotypical signature as well as a secondary, less strict phenotypical signature for sensitivity analyses.

The primary (cardiometabolic) phenotype included hypertension, above-cohort-median BMI and below-cohort-median hematocrit as well as chronic ischemic heart disease and absence of multiple myeloma, the latter to exclude likely AL cases. The secondary phenotype was limited to include hypertension, above-cohort-median BMI and below-cohort-median hematocrit.

### Outcome analysis

The primary endpoint was a composite of heart failure hospitalization or death. Mortality came from the Austrian Death Registry and hospitalizations from regional and national health records. We estimated survival with Kaplan-Meier curves as well as log-rank tests and modeled it with Cox proportional hazards adjusted for age, sex and past or present cancer. Proportional hazards were checked with scaled Schoenfeld residuals.

We first estimated the prognostic effect of Perugini grade (0, 1, ≥2). To confirm the grade 1 prognostic signal was not driven by referral or competing oncologic risk, we repeated the grade analysis after excluding amyloidosis referrals, after excluding cancer, and excluding both. We further assessed whether grade 1 patients reach or exceed the risk associated with grade ≥2 when restricted to the cardiometabolic phenotype.

### Unsupervised phenotype visualization

To place grade 1 within the full phenotype distribution, we projected the 1,243-variable feature set into two dimensions with UMAP (15). Variables missing in over 50% of patients (n=54) were dropped, continuous variables were median-imputed and standardized and binary variables mode-imputed. The embedding used Euclidean distance, 20 neighbors and a minimum distance of 0.1, and served qualitative illustration, with points colored by grade and phenotype-positive grade 1 patients highlighted.

### General statistical analysis

Continuous variables are reported as mean ± SD or median with IQR and categorical variables as counts with percentages. Groups were compared with t-tests or Mann-Whitney U tests for continuous variables and chi-square or Fisher’s exact tests for categorical variables. Analyses were performed in Python 3.9.5.

## Results

### Population characteristics

Of 9,170 patients referred for [^99m^Tc]Tc-DPD scintigraphy, 8,853 (96.5%) had no cardiac uptake, while grade 1 was present in 175 (1.9%) and grade ≥2 in 142 (1.6%) (**Supplemental Table S1**, **Figure 1**). Compared with grade 0, grade 1 patients were older (74.0 ± 9.5 vs 61.3 ± 15.7 years, p<0.0001) and more often male (55% vs 35%, p<0.0001). In contrast to the characteristically lean grade ≥2 group, grade 1 patients had a higher BMI (28.5 ± 5.4 vs 25.7 ± 5.4 kg/m², p<0.0001). Referrals for suspected cardiac amyloidosis accounted for 72/175 (41%) grade 1 and 116/142 (82%) grade ≥2 patients, the remainder being predominantly oncologic (**Supplemental Figure S1**). Grade-specific prevalence across all 1,243 clinical parameters is provided as a public resource (**Supplemental Table Prevalences**).

**Figure 1.**
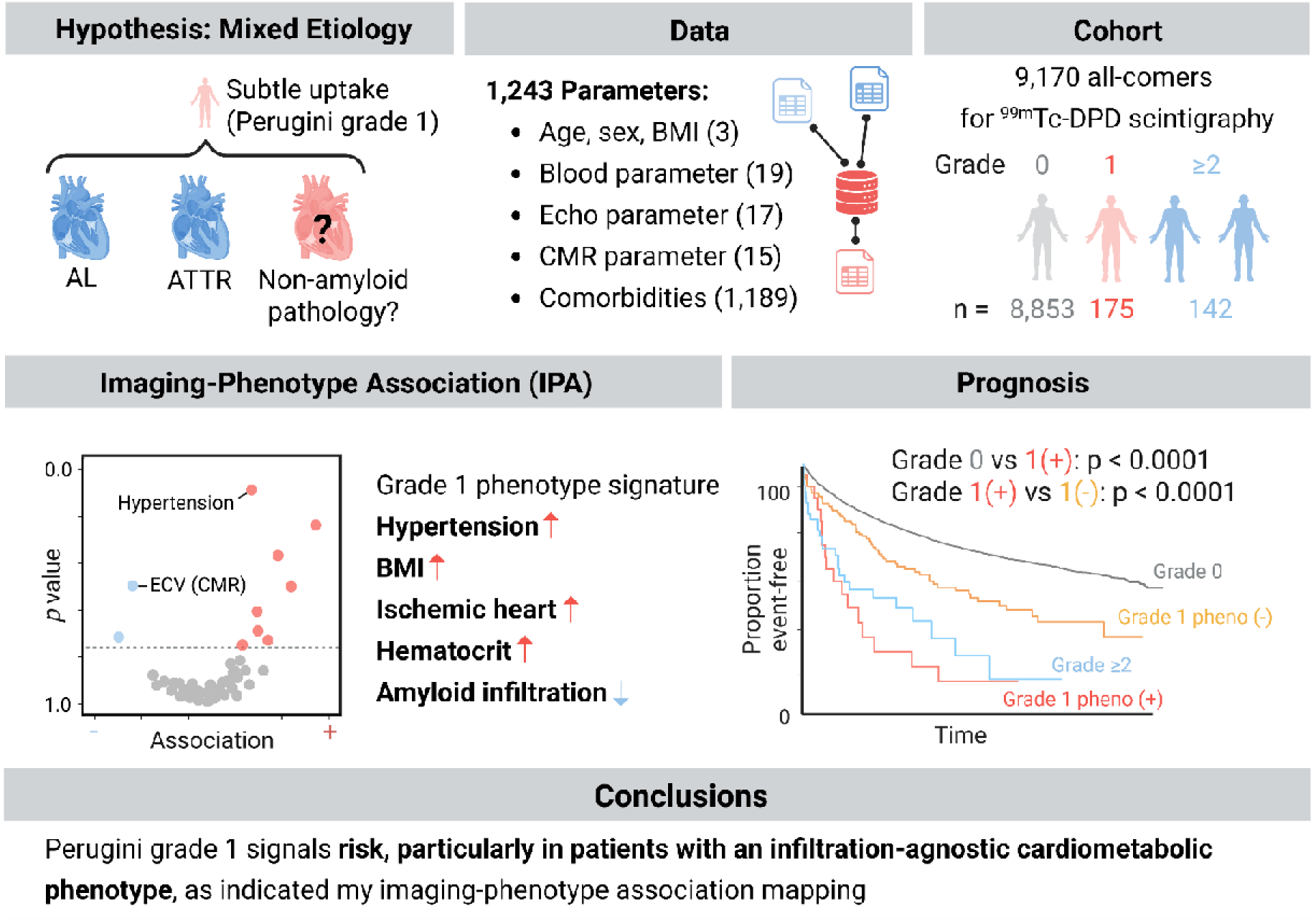
Study overview.

### Perugini grade predicts heart failure hospitalization and death

Cardiac uptake was associated with adverse outcomes (**Figure 2**). Grade ≥2 carried the worst prognosis (log-rank p<0.0001) and remained independently associated with the composite endpoint after adjustment for age, sex and cancer history (adjHR 1.70, 95% CI 1.35 to 2.13), consistent with infiltrative ATTR. Grade 1 also conferred excess risk over grade 0 (log-rank p<0.0001), smaller in magnitude but significant after adjustment (adjHR 1.30, 95% CI 1.07 to 1.59). Results were consistent for death and heart failure hospitalization analyzed separately, except for overall mortality after adjustment (**Supplemental Figures S2, S3**). The grade 1 signal persisted after excluding amyloidosis referrals, after excluding cancer, and after excluding both (p=0.0004, p<0.0001, p<0.0001), arguing against referral bias or competing oncologic risk as the explanation (**Supplemental Figure S4**).

**Figure 2.**
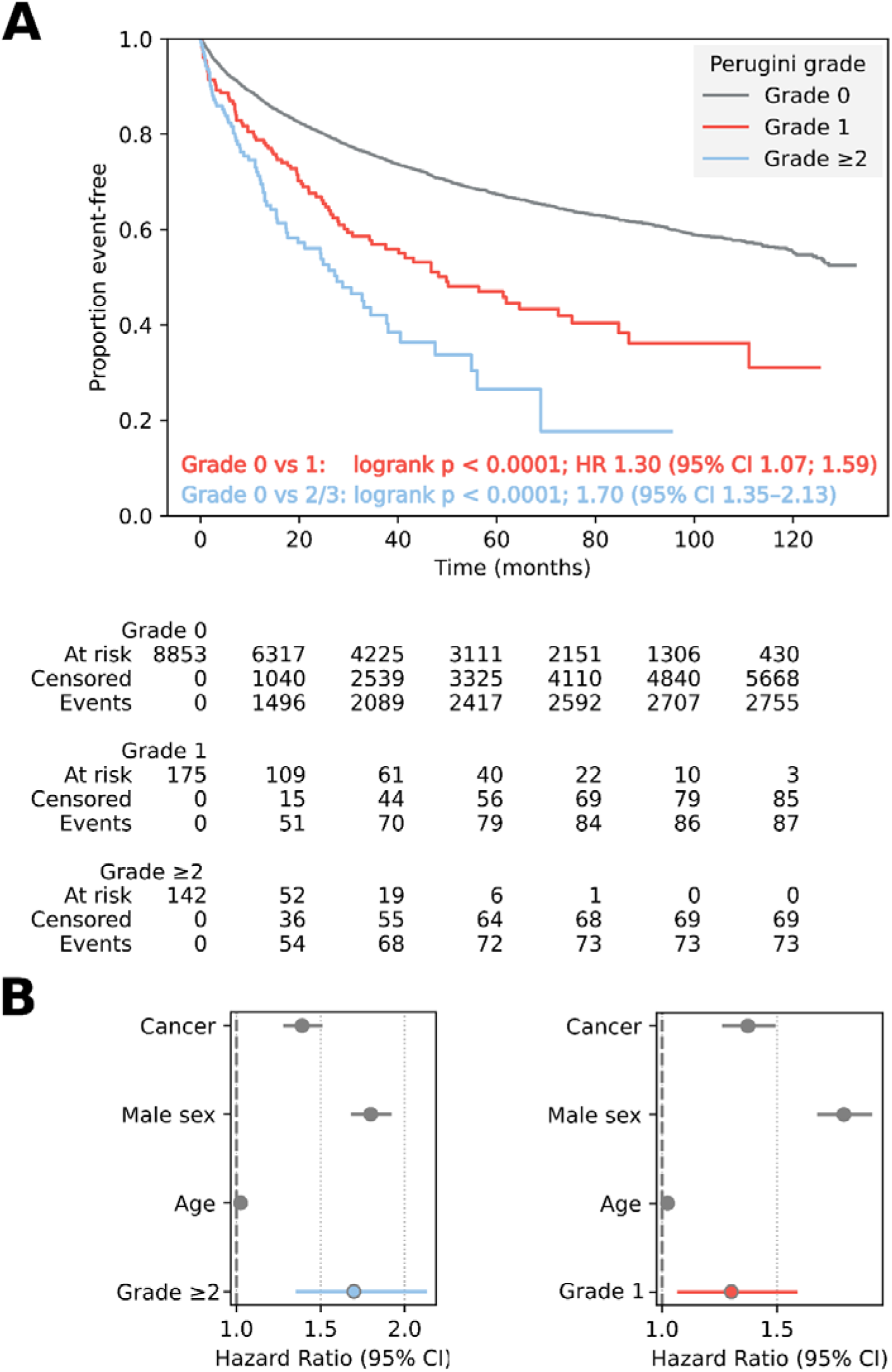
Prognostic association of Perugini grade ≥2 and grade 1 with death or heart failure hospitalization. Kaplan-Meier estimators (A) and Cox proportional hazards ratios (B).

### IPA recovers the infiltrative phenotype of grade **≥**2

As internal validation, comparing grade ≥2 with grade 0 reproduced the established ATTR phenotype (**Figure 3**). Neuropathy, atrial fibrillation, heart failure and cardiomyopathy were enriched (Bonferroni p<0.00014), with no significant negative associations. Continuous traits showed the expected infiltrative profile, with greater septal thickness, higher extracellular volume, atrial enlargement and elevated NT-proBNP and BUN, alongside lower eGFR, BMI and lipids (**Supplemental Table S2**). Recovering these known signatures confirms that IPA identifies clinically coherent phenotypes from routine data.

**Figure 3.**
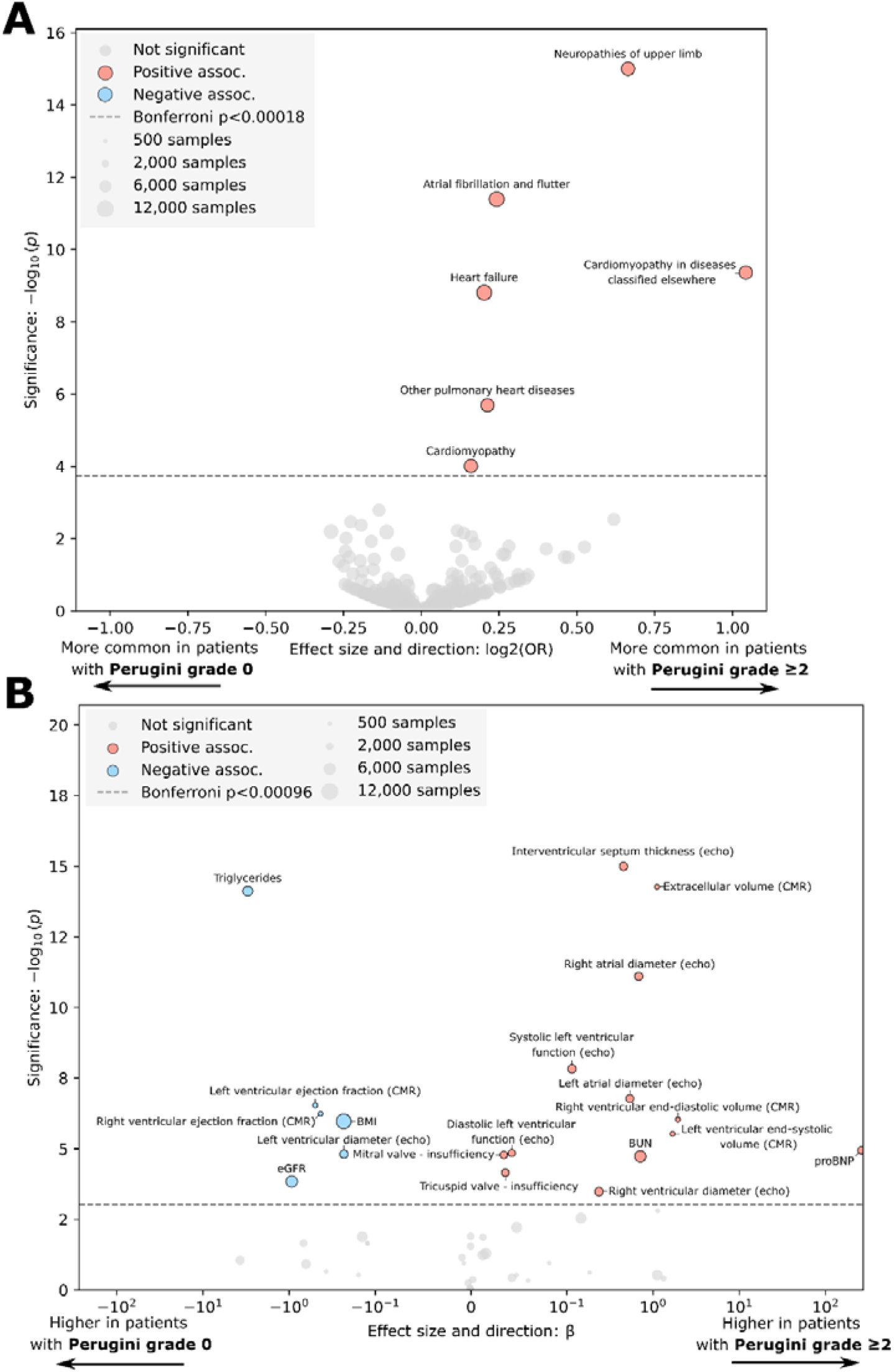
Association of Perugini grade ≥2 versus 0 with comorbidities (A) and continuous parameters (B). All positive associations were also identified by Benjamini-Hochberg FDR correction.

### Grade 1 marks a distinct cardiometabolic phenotype, not attenuated ATTR

Against grade 0, grade 1 showed a broad cardiovascular and systemic burden with smaller effect sizes compared to grade ≥2 (**Figure 4**). The parameters with the strongest associations were heart failure, atrial fibrillation or flutter and chronic ischemic heart disease, with further associations for hypertension, non-rheumatic mitral valve disease and chronic kidney disease as well as lower hemoglobin and hematocrit. Continuous traits showed elevated NT-proBNP and BUN, larger atrial diameters, more frequent mild tricuspid and mitral regurgitation, and reduced eGFR and total cholesterol (**Supplemental Table S3**). The overall pattern was consistent with a heterogeneous rather than a uniform amyloid state.

**Figure 4.**
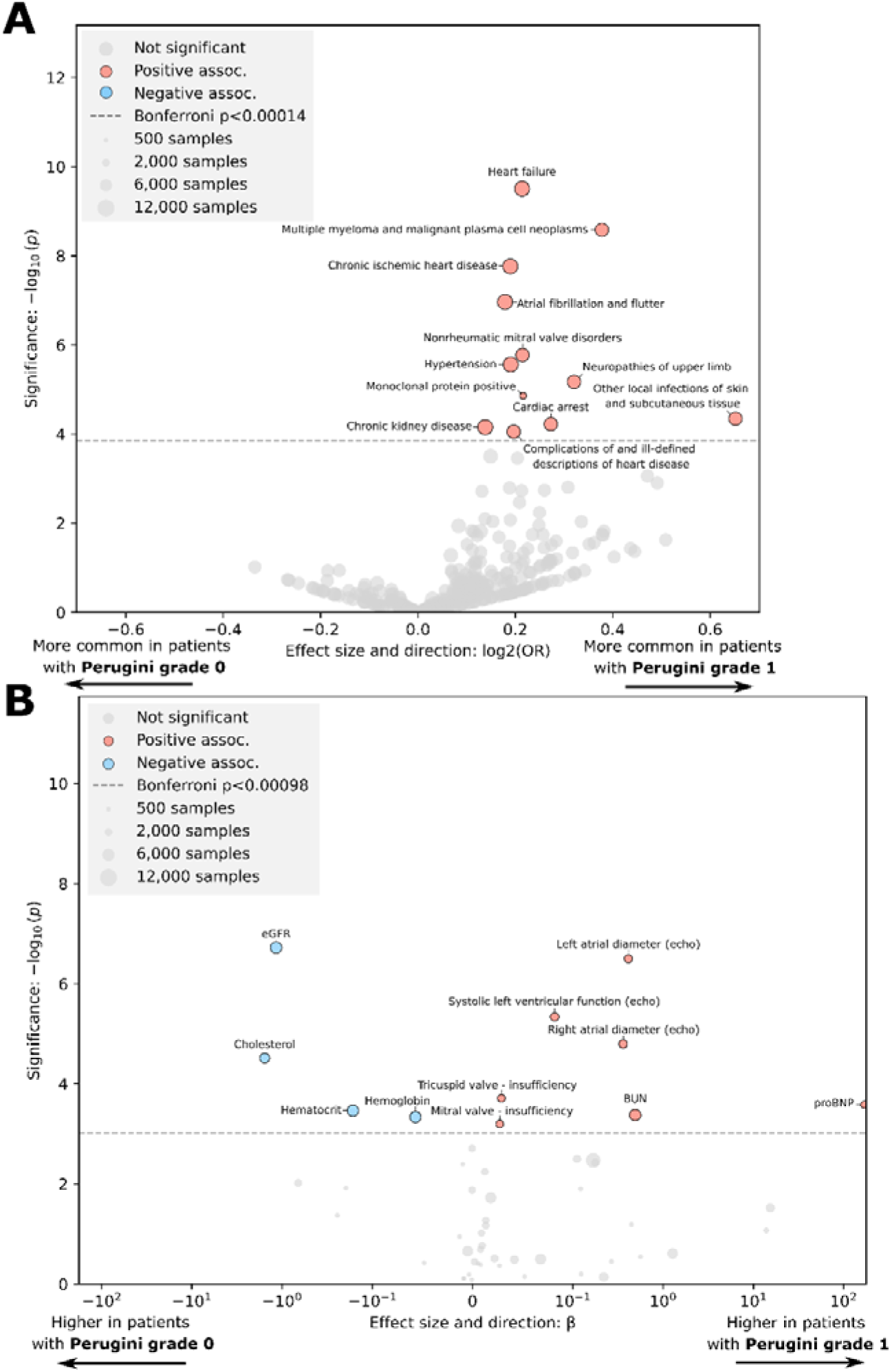
Association of Perugini grade 1 versus no cardiac uptake with comorbidities (A) and continuous parameters (B). All positive associations were also identified by Benjamini- Hochberg FDR correction.

Against grade ≥2, grade 1 diverged from the infiltrative phenotype (**Figure 5**). Grade 1 patients were more commonly affected by hypertension, had a higher BMI, lower hematocrit, lower hemoglobin and larger left ventricles, whereas septal thickness and extracellular volume were substantially lower and neuropathy was not enriched. A parallel positioning analysis classified each parameter by whether grade 1 resembled grade 0, resembled grade ≥2, or differed from both (**Supplemental Figure S6**). Cardiorenal markers, including elevated NT-proBNP, reduced eGFR, chronic kidney disease and chronic ischemic heart disease, resembled grade ≥2. Septal thickness and extracellular volume were intermediate but far closer to grade 0, arguing against meaningful infiltration. Hypertension, BMI and anemia exceeded both groups, identifying features that cannot represent an attenuated form of amyloid and instead point to a distinct cardiometabolic profile.

**Figure 5.**
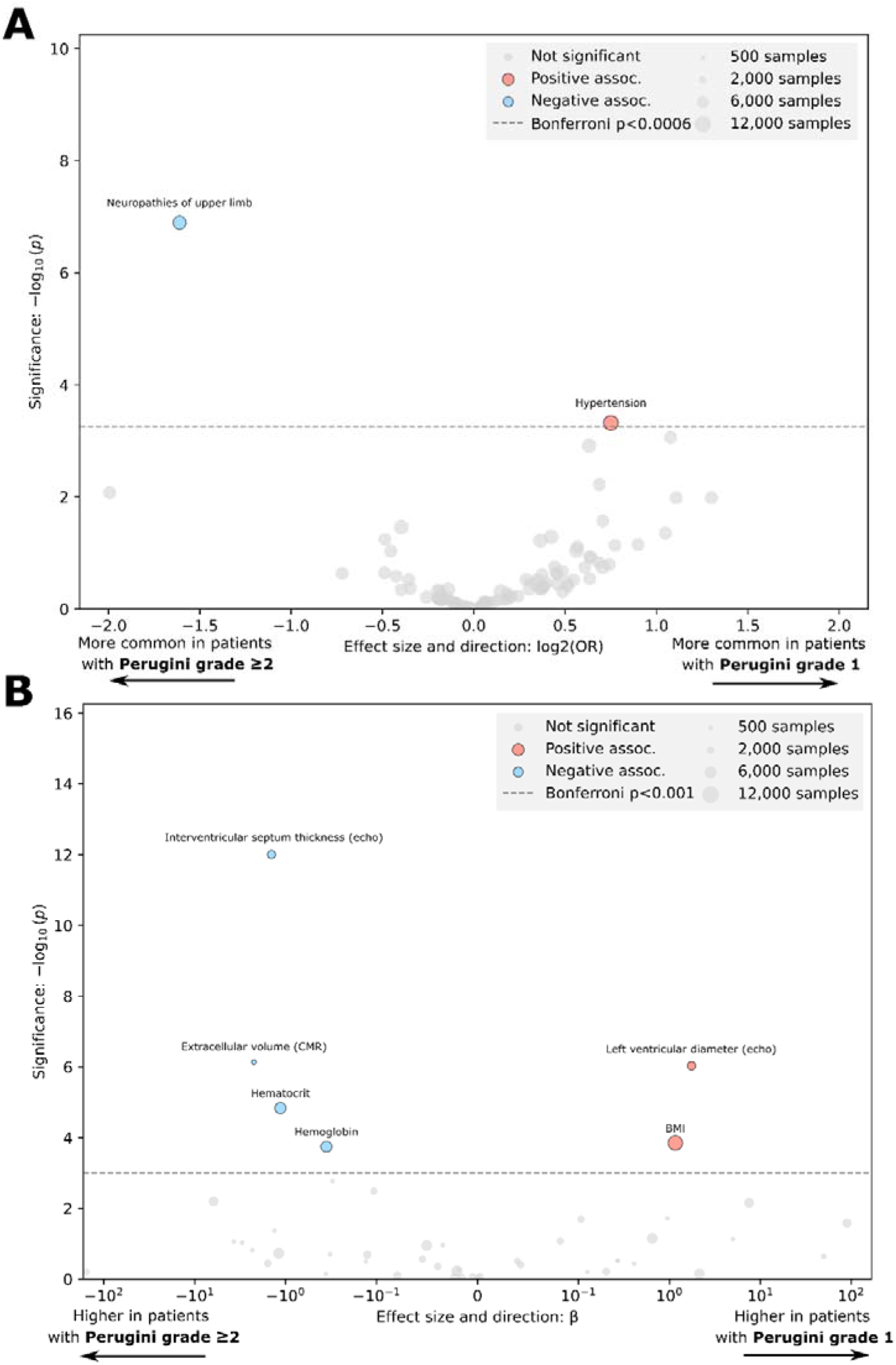
Association of Perugini grade 1 versus ≥2 for comorbidities (A) and continuous parameters (B). All positive associations were also identified by Benjamini-Hochberg FDR correction.

### Definition and structure of the cardiometabolic phenotype

From the parameters that distinguished grade 1 from grade 0 or grade ≥2, we constructed a cardiometabolic phenotype, excluding intermediate parameters and those available in less than 50% of patients. The primary phenotype required hypertension, above-median BMI and below-median hematocrit as well as chronic ischemic heart disease and absence of multiple myeloma, and was present in 23 of 175 (13%) grade 1 patients. A less strict phenotype definition required hypertension, above-median BMI and below-median hematocrit, present in 54 of 175 (31%). This phenotype is intended to enrich for non-amyloid cardiometabolic disease but, given sparse CMR coverage and the absence of biopsy, cannot exclude coexisting ATTR or AL.

Projecting the 1,243-variable phenome using UMAP placed the three grades on a shared, overlapping distribution with visible graded structure (**Figure 6**). Grade 1 did not occupy a single region but was distributed between grade 0 and the grade ≥2 enriched area, consistent with a mixture. Phenotype-positive grade 1 patients did not scatter randomly but concentrated in a confined region, indicating that this hypertensive, higher-BMI, lower-hematocrit profile is a coherent and recurring pattern rather than an arbitrary definition.

**Figure 6.**
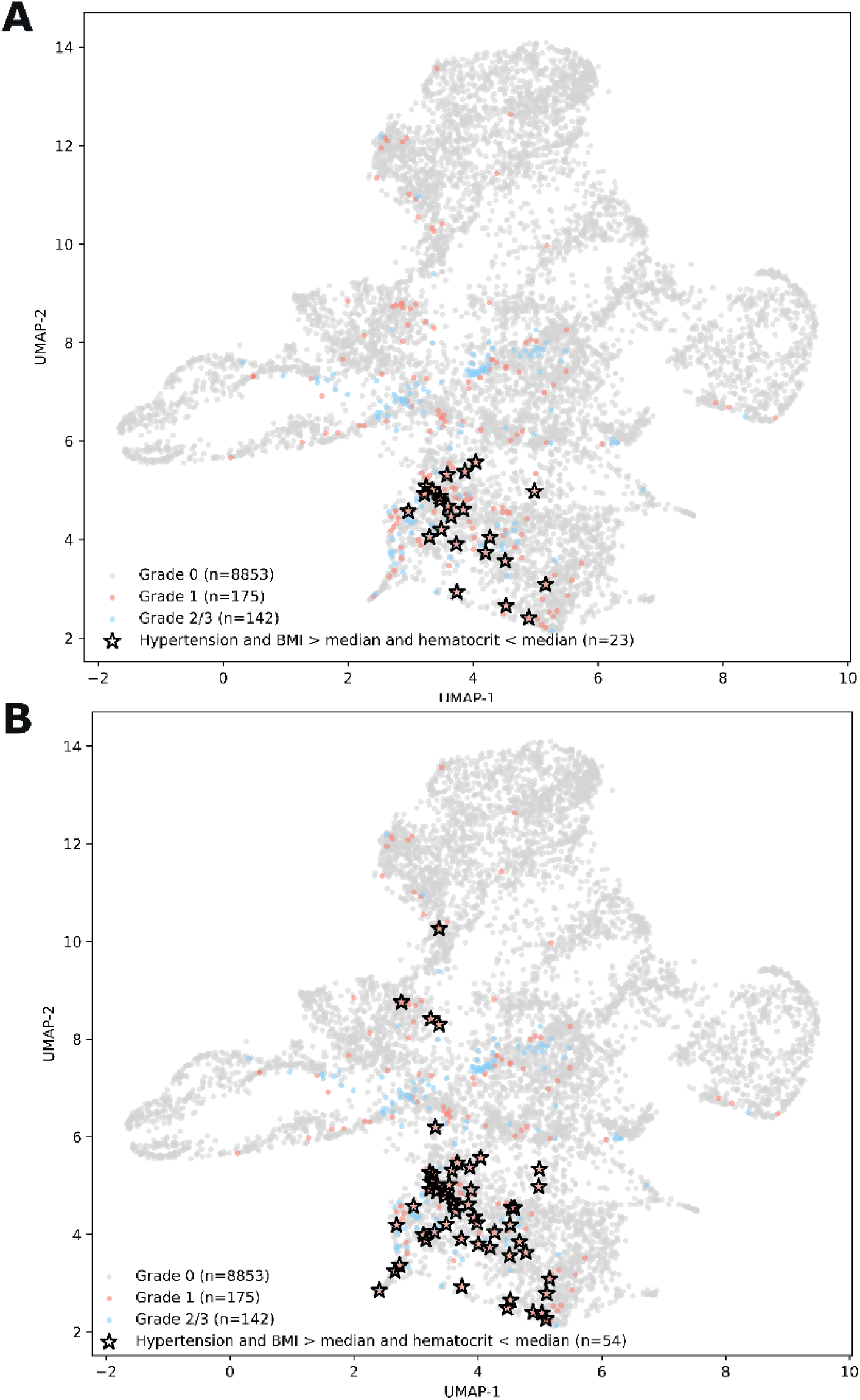
Unsupervised phenome visualization with annotations for the defined primary phenotype signature (**A**) and the secondary, less strict phenotype (**B**).

### The cardiometabolic phenotype drives grade 1 prognosis

To test whether grade 1 prognosis is carried by the cardiometabolic phenotype, we stratified patients with grade 1 into phenotype-negative and phenotype-positive (**Figure 7**). Phenotype-positive grade 1 patients had markedly worse outcomes than phenotype-negative patients (adjHR 1.98 (95% CI 1.18-3.34), p=0.01) and reached or exceeded the risk of grade ≥2 (adjHR 2.46 (95% CI 1.53-3.95), p=0.0002 vs adjHR 1.76 (95% CI 1.22-2.54), p=0.0026) (**Figure 7B**). The more inclusive sensitivity phenotype showed a consistent effect (**Supplementary Figure S7**). These results indicate that grade 1 prognosis is driven substantially by comorbid cardiometabolic disease, while a minority of grade 1 patients with ATTR or AL exist within this group.

**Figure 7.**
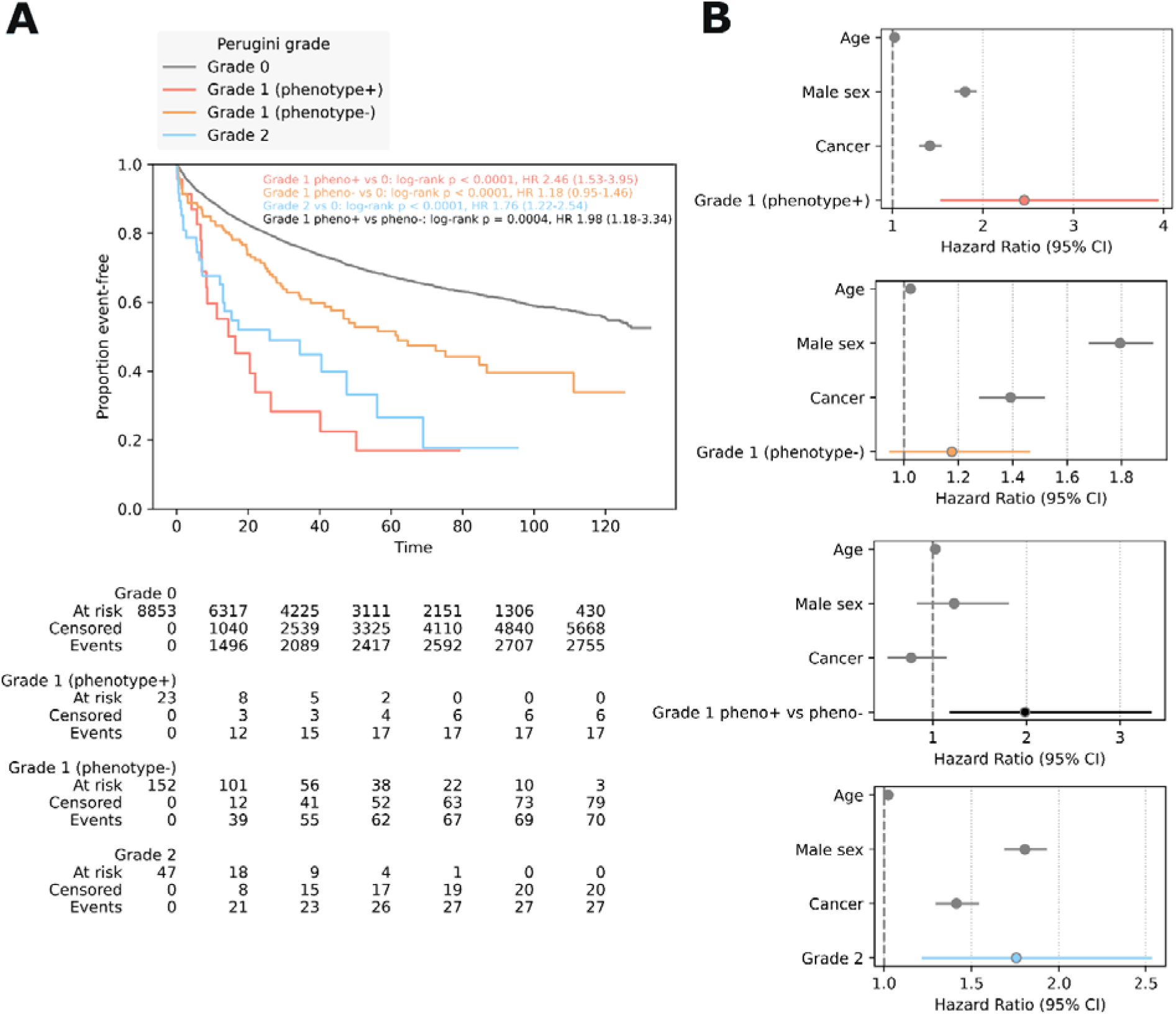
Prognostic association of grade 1 with (+) and without (-) the cardiometabolic phenotype (hypertension, high BMI, low hematocrit, chronic ischemic heart disease, no multiple myeloma), shown via Kaplan Meier estimators (A) and multivariable hazard plots (B).

### Exploratory SPECT/CT subgroup

SPECT/CT was available in 24 grade 1 patients, showing myocardial localization in 11 (46%) and predominant blood-pool activity in 13 (54%) patients. Outcomes did not differ by localization (HR 0.99, 95% CI 0.27 to 3.70), however, this subgroup was small and likely underpowered, so the comparison is hypothesis-generating and cannot establish equivalence (**Supplemental Figure S5, Table S5**). Nonetheless, it is consistent with the planar phenotype, rather than indicating that tracer localization alone relates to prognosis.

## Discussion

In a large all-comer cohort undergoing bone scintigraphy, we used an imaging-phenotype association framework to characterize the clinical meaning of Perugini grade 1 and tested whether its prognosis is driven by amyloid or by the comorbidity it accompanies. Three findings emerged. Grade 1 marks a distinct cardiometabolic phenotype rather than attenuated ATTR. This phenotype, defined on routine comorbidity and laboratory markers, identifies grade 1 patients whose risk reaches or exceeds that of grade ≥2. The effect is specific to grade 1 rather than a generic comorbidity effect, supporting the interpretation that non-infiltrative cardiometabolic disease substantially impacts the prognosis of patients with Perugini grade 1. As validation, the grade ≥2 versus grade 0 comparison recovered the established ATTR phenotype, with increased wall thickness and extracellular volume, atrial enlargement, peripheral neuropathy and elevated natriuretic peptides (16,17). Reproducing these known signatures from routine data shows that IPA identifies clinically coherent imaging phenotypes and provides an analytical framework for the analysis of the more ambiguous grade 1 phenotype.

Grade 1 did not resemble a faint version of the amyloid phenotype. Although grade 1 patients shared cardiorenal stress markers with grade ≥2, including higher NT-proBNP and BUN and lower eGFR, these were smaller in magnitude and occurred without the structural hallmarks of infiltration. Septal thickness and extracellular volume were similar to those of patients with grade 0 than grade ≥2 uptake. Conversely, hypertension and BMI and anemia exceeded both grades, features that cannot represent an attenuated form of ATTR or AL and instead point to a non-infiltrative cardiometabolic process. The unsupervised phenome analysis supported this, placing grade 1 across a shared distribution while the cardiometabolic phenotype-positive subset concentrated in a confined region, indicating a coherent, recurring pattern rather than an arbitrary threshold.

The cardiometabolic phenotype had important implications for prognosis. Within grade 1, the phenotype identified patients whose outcomes matched or exceeded those of grade ≥2, despite the absence of infiltrative features. We emphasize that a subset of grade 1 patients certainly harbor ATTR or AL, evidenced here by monoclonal protein and multiple myeloma enrichment. The identified phenotype is likely to enrich for non-amyloid disease rather than to rule amyloid out, which the sparse availability of CMR and absent biopsy data do not permit. These findings carry practical implications. Grade 1 on planar scintigraphy should be read neither as an artifact nor as presumptive amyloid, but as a marker that frequently accompanies prognostically meaningful cardiometabolic disease. In patients with the cardiometabolic phenotype and no other amyloid red flags, attention to hypertension, renal function, ischemic heart disease and heart failure management may matter as much as amyloid workup, whereas disproportionate wall thickening, extracellular volume expansion or neuropathy should still prompt confirmatory amyloid-directed evaluation. IPA offers a scalable way to support such phenotype-based interpretation, as quantitative cardiac imaging is more widely integrated into routine care. As part of this work, we provide grade-specific prevalence across more than 1,200 ICD-10 phenotypes as a public resource to support hypothesis generation and comparative phenotyping (**Supplementary Table Prevalences**).

All findings in this study pertain to the planar Perugini grade. This is not a departure from the Perugini score itself, which was defined on planar images (13), but it does depart from current practice recommendations for the diagnosis of ATTR cardiac amyloidosis, which require SPECT to establish that cardiac activity is myocardial before any positive grade is assigned (13,18,19). We regard this as the point of the analysis rather than a defect of it. The overwhelming majority of bone scintigraphy performed worldwide is acquired for oncologic indications without thoracic SPECT, yet a planar grade is still assigned and still reaches the referring clinician. Patients labeled planar grade 1 in that setting are guideline-orphaned, and our data indicate that their prognosis is legible from routine comorbidity rather than requiring amyloid-specific imaging they will not receive.

Our findings extend prior work. Earlier all-comer analyses established that grade 1 carries excess risk over grade 0 but did not explain its source (10,11). SPECT/CT series showed that most grade 1 uptake is blood-pool rather than myocardial and described the associated comorbidity load in small samples (6–9). A recent variant-carrier study addressed diagnostic conversion to ATTR rather than prognosis (20). In an overlapping cohort from this center with external validation at an independent institution, we previously demonstrated that structured EHR data predicts amyloid-suggestive grade ≥2 uptake with high diagnostic performance and can guide referral for confirmatory scintigraphy (25). While the previous work addresses which patients should be scanned, the present analysis addresses how to interpret the scan when the result is equivocal, covering a distinct clinical gap. The present study further moves beyond all previous studies by systematically characterizing grade 1 across more than 1,200 phenotypes at scale and by linking a defined clinical phenotype to its prognosis, offering a mechanism-agnostic explanation for why a largely non-myocardial signal predicts outcomes.

This study has several limitations. Visual grading of planar uptake is subject to interreader variability and possible misclassification in borderline cases, although multi-reader consensus provides the most rigorous feasible planar assessment (21–23). The SPECT/CT subgroup was small and underpowered, so the absence of an outcome difference between myocardial and blood-pool localization is hypothesis-generating and does not establish equivalence. Confirmatory imaging and biopsy were unavailable in most patients, and CMR-derived extracellular volume was sparse, which limits direct exclusion of infiltration in the phenotype-positive subgroup. The phenotype was derived in this cohort and, although fixed before any survival testing to limit optimization, requires external validation. Routine EHR data carried missingness that was imputed and may weaken associations, and monoclonal protein was assessed only in a subset, limiting inference about AL.

## Conclusion

Perugini grade 1 on [^99m^Tc]Tc-DPD scintigraphy is not a uniform early amyloid signal nor a benign artifact but a marker of a heterogeneous, predominantly non-infiltrative state carrying clinically meaningful risk. A simple cardiometabolic phenotype defined on routine comorbidity and laboratory data identifies grade 1 patients whose prognosis matches or exceeds that of amyloid-suggestive grade ≥2 uptake, indicating that much of the adverse outcome associated with grade 1 is driven by comorbid cardiometabolic disease rather than amyloid burden, even as a minority of patients harbor ATTR or AL. These findings support reinterpreting grade 1 as a mixed-etiology phenotype and prioritizing cardiorenal assessment alongside selective amyloid workup. The IPA framework provides a scalable approach to refining the interpretation of cardiovascular imaging biomarkers.

## Statements and Declarations

### Author Contributions

CPS conceived and designed the study. CPS, JN, KM, KK, MP, RR, FH, CN and RC collected and annotated the data. CPS developed the related code and performed the statistical analysis. CPS wrote the manuscript and created the figures. JN, DK and RC provided feedback regarding clinical interpretation. CH and MH provided supervision. All authors critically reviewed the manuscript and provided feedback.

### Data Availability

The current ethical approval of this study does not permit the sharing of study data. If you would like to access the data, please contact the corresponding author.

### Disclaimer

Automated writing software, including Grammarly and Claude were used to support the writing of this manuscript. All results and conclusions reflect the authors’ own.

### Ethics

This study was performed in line with the principles of the Declaration of Helsinki. Approval was obtained from the Ethics Committee of the Medical University of Vienna (No. 2278/2024). The institutional review board waived the requirement for informed consent owing to the retrospective design of the study.

